# Differential gene expression in scrub typhus compared to other acute febrile infections by bioinformatic approaches

**DOI:** 10.1101/2020.12.21.20248609

**Authors:** Manisha Mandal, Shyamapada Mandal

## Abstract

Scrub typhus (ST), caused with the infection of *Orientia tsutsugamushi*, without eschar, is a febrile illness that mimics malaria (ML), dengue (DG), and other rickettsioses such as murine typhus (MT). Comparative analysis of microarray gene expression profiles of GSE16463 dataset, from *O. tsutsugamushi* infected monocytes, was performed to identify transcriptional signatures in ST discriminated from other acute febrile infections, accompanied by functional pathways and enrichment analysis in disease pathogenesis. A unique 31 ST-associated signature genes obtained in this study could help distinguish ST from other febrile illnesses DG, ML and MT. The functional pathways significantly enriched in ST disease group included translocation of ZAP-70 to immunological synapse, and phosphorylation of CD3 and TCR zeta chains, involving PTPN22 and CD3G genes, which could further help in the understanding of molecular pathophysiology of ST and discovering novel drug targets as well as vaccine developments.

## Introduction

Concerning the acute febrile infection (AFI) development of improved diagnostics, particularly the molecular diagnostic intervention with therapeutic potential, is an urgent need for infection control and treatment management. Scrub typhus (ST), which is caused with the infection of a bacterial pathogen *Orientia tsutsugamushi* (vectored by larval trombiculid mites *Leptotrombidium deliense*), presents as a febrile illness with the severity ranging from mild (no organ dysfunction), to moderate (with one organ dysfunction) to severe/lethal (two or more organ dysfunction: renal dysfunction, pulmonary dysfunction, cardiovascular dysfunction, or central nervous system dysfunction) (Astrup et al., 2014). The ST along with murine typhus (MT) is as prevalent as malaria (ML) and dengue (DG), displaying symptoms that include fever and chill, headache, body aches, and sometimes eschar or rash (at initial stages), and causes substantial morbidity and mortality worldwide (CDC, 2020; Diaz et al., 2018). The ST (in absence of distinctive eschar at the inoculation site) mimics the other febrile diseases MT, DG and ML, and thus due to the non-specific clinical presentation distinction of ST is very difficult from MT, ML and DG in geographical areas where these diseases prevail, and in addition the lack of reliable point-of-care (sensitive and specific) diagnostic tests, diagnosing ST is difficult. Therefore, early diagnosis of ST using effective molecular-based tools is crucial in order to provide prompt antimicrobial treatment to prevent the disease complications and reduce the mortality rate (Astrup et al., 2014). Bioinformatics approaches have been found useful in identifying signature genes as well as predicting diseases outcome. Herein we investigated the molecular response that shows up distinct to these four categories of infection (ST, from MT, ML and DG) with overlapping signs and symptoms using GSE1646 dataset, to track the transcriptome, to gauge the differentially expressed genes (DEGs), investigate underlying molecular interactions between DEGs, functional enrichment and significant pathways associated with the DEGs, protein-protein interaction (PPI) network of common DEGs, to identify the genes as predictive biomarkers as a basis for a targeted therapy that can help guide more accurate therapeutic clinical studies and, in the future, to provide treatment decisions. Detecting ST-associated signature genes have been reported to contribute in revealing the disease pathogenesis enabling the diagnosis, discovery of novel drug targets for ST (Tantibhedhyangkul et al., 2011), and vaccine development as well (Valbuena and Walker 2013). Therefore, analysis of the DEGs in ST, along with other infectious febrile illnesses (ML, DG and MT), compared to healthy individuals might be an expedient tool in determining the biomarkers of clinical outcome of infections to facilitate the assessment of the need for clinical interventions.

## Methods

The dataset GSE1646 containing genome-wide expression in peripheral blood mononuclear cells from patients with ST (n=4; with fever duration of 3 – 20 days from cases of 19 – 56 years of age), DG (n=7; with fever duration of 1 – 4 days from cases of 18 – 70 years), MT (n=7; with fever duration of 6 – 21 days from cases of 23 – 65 years), and ML (n= 4; with fever duration of 3 – 7 days from cases of 24 – 34 years), compared to two healthy controls (HC), deposited by Tantibhedhyangkul et al. (2011), was downloaded from the GEO (gene expression omnibus) database (www.ncbi.nlm.nih.gov/geo/) and extracted with GEO2R tools (http://www.ncbi.nlm.nih.gov/geo/geo2r/) in order to perform comparisons with GEOquery, limma R, Biobase packages from the Bioconductor project (Barrett et al. 2013; Huber et al. 2015). The raw microarray data obtained was processed for “NA” filtering and normalized applying “normalizeBetweenArrays” in limma, using quantile method followed by fitting with a linear model and comparison among disease groups: ST, DG, MT and ML, using empirical Bayes moderated t test, in limma, to generate log2FoldChange (log2FC) differential expression of ST, DG, MT and ML, the P values and FDR (False Discovery Rate) adjusted P value (adj.P.value) with cutoff criterion for DEGs set as P value <0.05 and |log2FC| >1 (Mandal and Mandal 2020).

The regulatory network of the DEGs was built using cytoscape (https://cytoscape.org/), version 3.8.2 (Shannon et al. 2003). The PPI network was created using STRING (Search Tool for the Retrieval of Interacting Genes) (http://string-db.org/cgi/input.pl; version 1.6.0) App of the cytoscape [16], version 3.8.2 (https://cytoscape.org/) (Shannon et al. 2003). The important hub genes within the PPI network were identified using the cytoscape plugin CytoHubba, version 0.1, ranked by higher scores as per maximal clique centrality (MCC) method, amongst several topological algorithms (Chin et al. 2014). The significant modules containing cluster of densely connected regions within the PPI network were explored using the MCODE (molecular complex detection) version 2.0.0 cytoscape plugin, with threshold values of degree 2, node score 0.2, k-core 2 and maximum depth 100 (Bader and Hogue 2003). The DEGs were subjected to GO (gene ontology) enrichment analysis to explore the functional roles in terms of biological processes, cellular components and molecular functions using STRING Enrichment App of the cytoscape software, with threshold P value <0.05 (Doncheva et al. 2019). The DEGs were also examined to study the statistical enrichment in Reactome (https://reactome.org) pathways, using the STRING Enrichment App.

## Results and Discussion

We performed a comparative analysis of microarray gene expression profiles from GSE16463 dataset using bioinformatics approaches, in connection with *O. tsutsugamushi* infection in human monocytes to identify the transcriptional signatures from gene interaction networks in ST discriminated from other acute febrile illnesses including DG, ML, MT accompanied by gene ontology and functional pathway enrichment analysis in disease pathogenesis. The ST, DG, MT and ML gene expression microarray signal intensity downloaded from NCBI GEO dataset GSE16463 before and after normalization have been indicated in Figure 1A and Figure 1B. Analysis of normalized expression data using GEO2R yielded 96 DEGs indicated as transcript IDs for the 24 samples of ST, DG, MT, ML and HC with a heatmap displaying hierarchical structural analysis among the samples and transcripts as well (Figure 1C). The empirical Bayes moderated t distribution for the log2FC of ST, DG, MT, ML, HC compared to each other, average Expression (AveExpr), F-statistics (F), the P values, and adj.P.values (Figure 1D).

**Figure 1:**
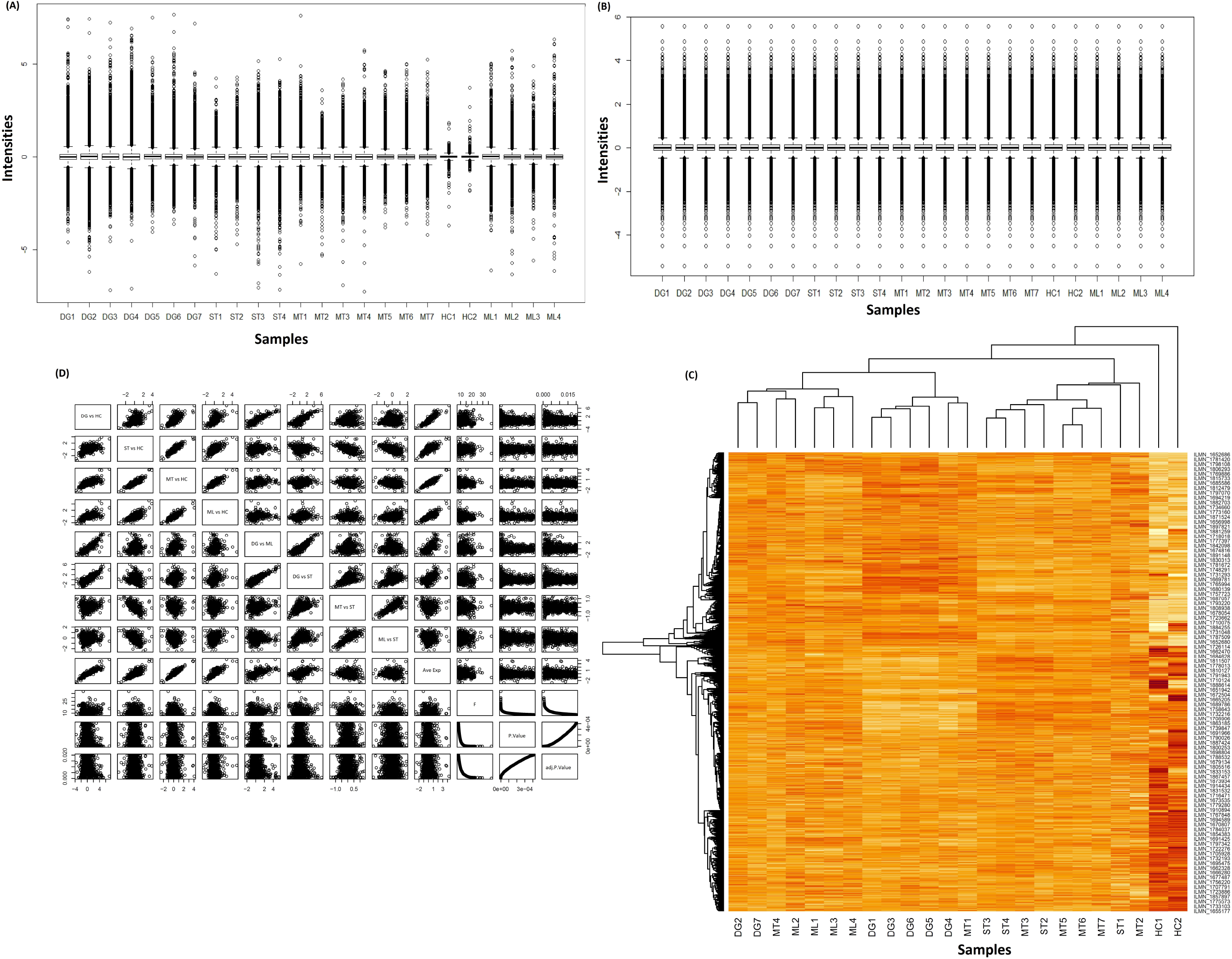
Microarray expression of RNA signal intensity from peripheral blood mononuclear cells in scrub typhus (ST; n=4), dengue (DG; n=7), murine typhus (MT; n=7), malaria (ML; n= 4) and healthy controls (HC; n=2): (A) before and (B) after normalization; (C) heatmap of normalized microarrays in ST, DG, MT, ML and HC; (D) empirical Bayes moderated t distribution.

The upregulated and downregulated DEGs among ST, DG, ML, MT compared to HC groups have been represented with volcano plots in Figure 2. The scrub typhus regulatory networks consisted of 150 genes, of which 65 were upregulated and 85 downregulated with log2FC ranging from 1.006819243 (for PTPN22) to 3.87069874 by (for ANKRD22) and from -1.0598985 (for CBX7) to -2.288928002 (for GAL3ST4), respectively (Figure 2A). Biological regulation, cell surface receptor signalling pathway, immune response and its regulation, cytokine-mediated signalling pathway, lymphocyte activation, protein localization to membrane were significantly overrepresented among the differentially expressed genes in ST disease (Table 1). Earlier Park et al. (2018) reported 70 upregulated proteins and 94 downregulated proteins ensuing with the *O. tsutsugamushi* infection compared to normal subjects, with the expression of proteins involved in immune responses, particularly the acute phase response signaling, complement system, LXR/RXR activation, FXR/RXR activation and coagulation system upregulation. The most significantly enriched reactome pathways in ST group, from our study, were immune system, cytokine signalling, and immunoregulatory interactions between a lymphoid and a non-lymphoid cell (Table 1). The pathways enriched in differentially expressed genes in scrub typhus included surface proteins and adhesins, DNA replication, secreted effector protein, RAGE (rickettsial-amplified genetic element), and metabolic pathway, with upregulation of IFNB1 (interferon beta 1) gene, genes involved in regulating the type-I interferon response: IRF9 and STAT1/2, and interferon-stimulated genes (IFIT, OAS1) as well as proinflammatory chemokine (CXCL10, CXCL11) and cytokine receptor (IL13RA2, IL7R, IL15RA, IL3RA) genes (Mika-Gospodorz et al., 2020).

**Table 1.**
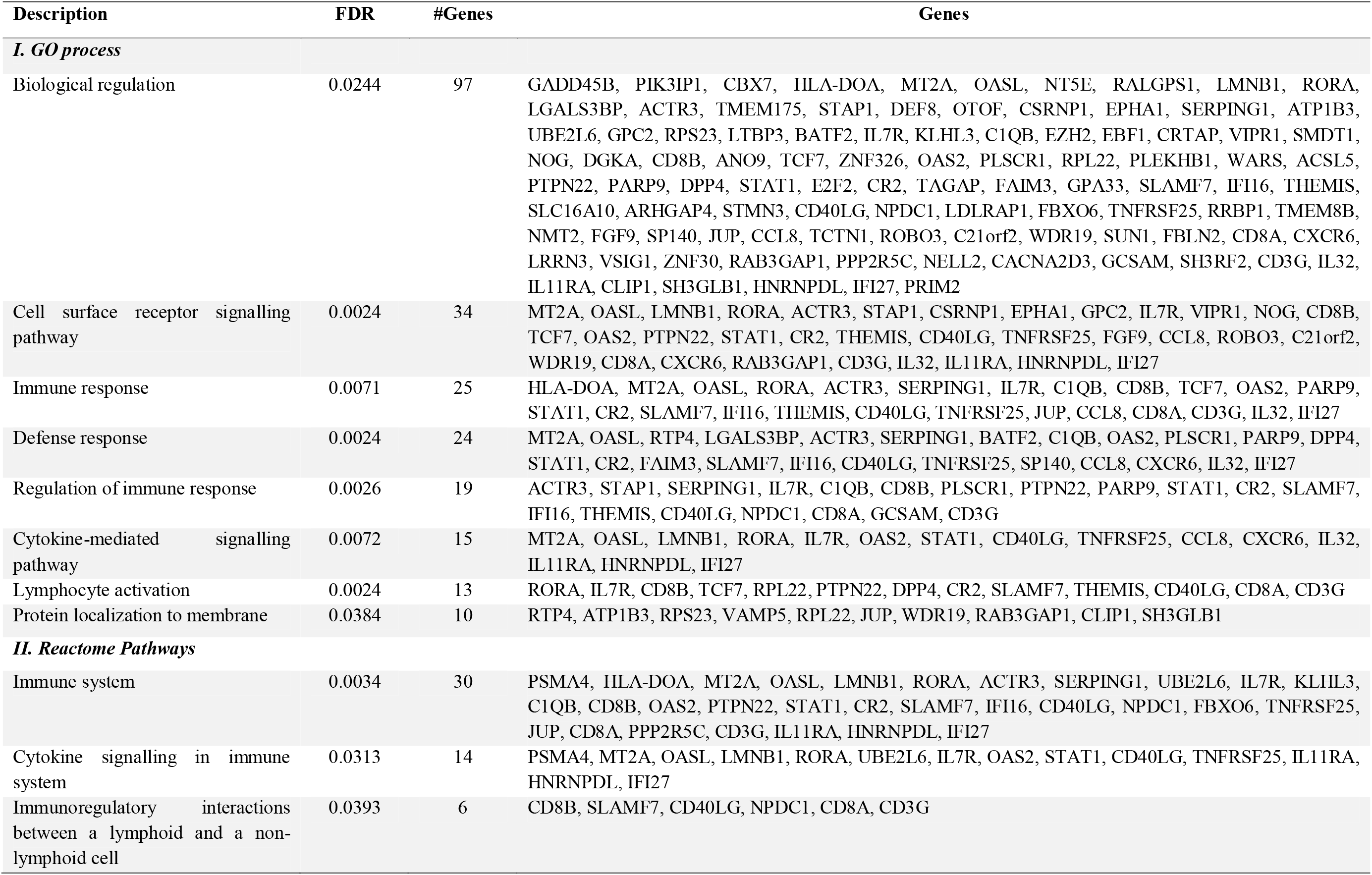
Enriched GO processes and reactome pathways for DEGs in scrub typhus.

**Figure 2:**
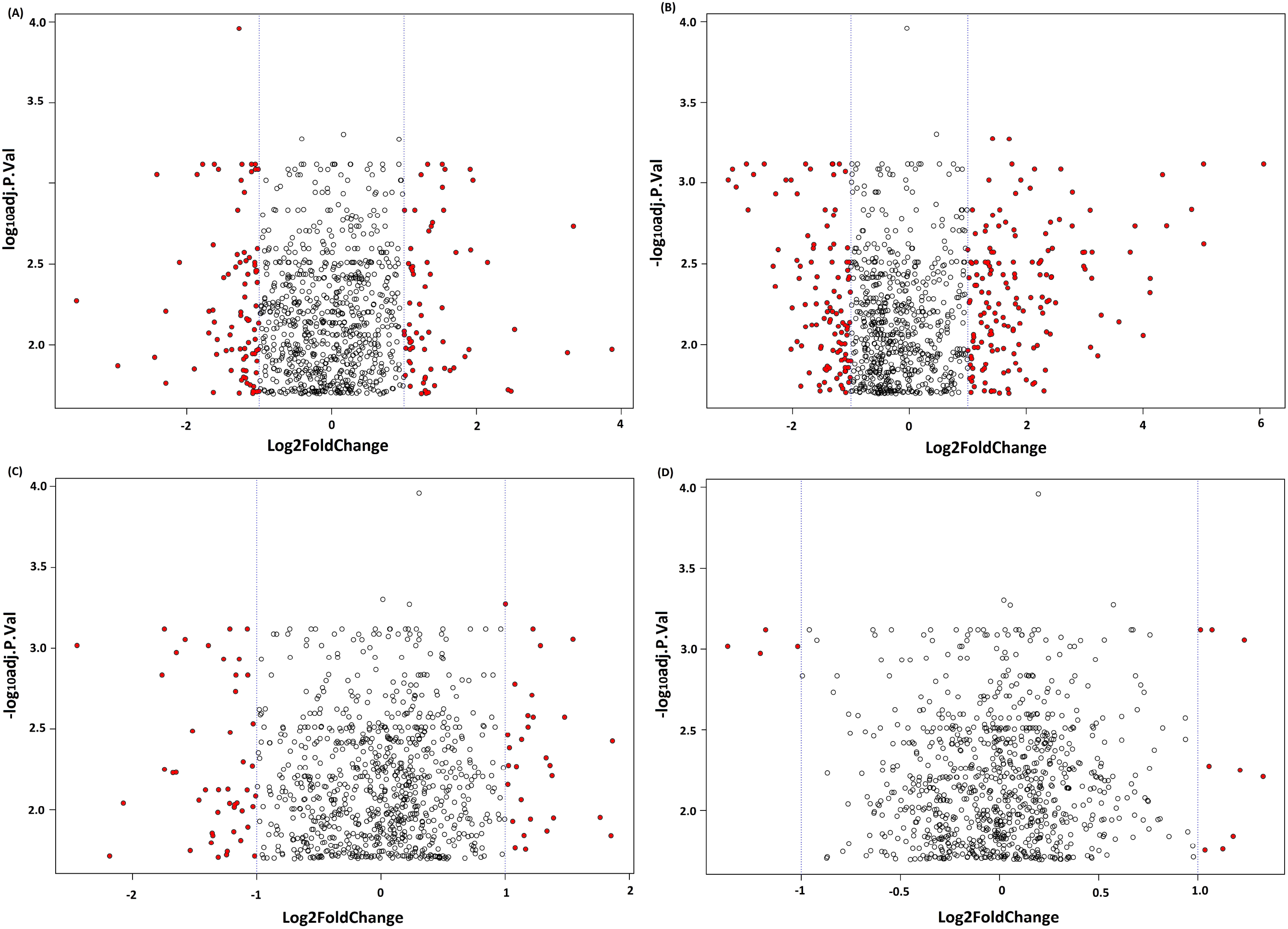
Volcano plots representing upregulated and downregulated expression of genes in (A) scrub typhus, (B) dengue, (C) malaria, (D) murine typhus, ccompared to healthy controls.

The dengue regulatory network (Figure 2B), consisted of 297 genes with 138 upregulated that exhibited log2FC ranging from 1.011207 (for ARSB) to 7.201346 by (for IFI27), and 159 downregulated genes for which log2FC ranged from -1.00412 (for IL32) to -0.31158 by (for LRRN3). Immune system and its innate type along with gamma interferon signalling, signalling by interleukins, ISG15 and OAS antiviral mechanism, L13a-mediated translational silencing of ceruloplasmin expression, negative regulators of DDX58/IFIH1 signalling, TRAF3-dependent IRF activation pathway were among the overstimulated reactome pathways in DG-associated genes (Table 2).

**Table 2.**
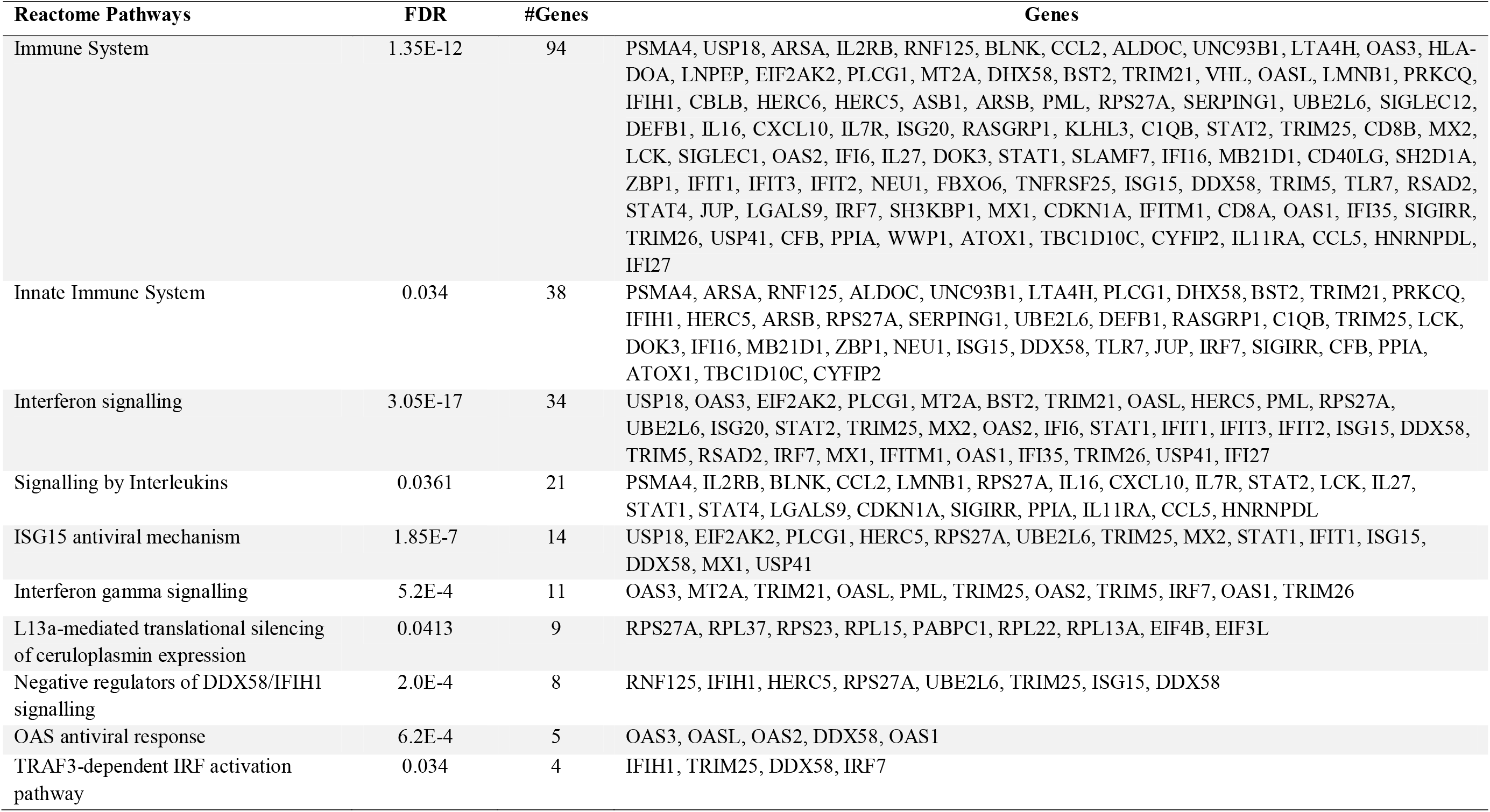
Enriched reactome pathways for DEGs in dengue fever.

Among a total of 53 genes within the ML regulatory network (Figure 2C), 28 were upregulated and 25 downregulated displaying log2FC ranging from 1.000882 (for ZBED2) to 2.737276 (for IFI27) and from - 1.01912 (for RNF125) to -2.42052 (for LOC649841), respectively. The immune system involving cytokine signalling was the most significant reactome pathway in ML group associated with genes including IFI27, RNF125, MT2A, TRIM21, LMNB1, SERPING1, UBE2L6, CXCL10, KLHL3, C1QB, SLAMF7, CD40LG, FBXO6, TNFRSF25, LGMN, JUP, IFI35, CFB, WWP1, ATOX1, and IL11RA (Table 3). In a network-based approach, 27 top ranked candidate genes were enriched in malaria patients through pathway analysis, with the expression of pyruvate metabolism, TCA cycle, integrin and vascular wall cell surface interactions, along with platelet adhesion to exposed collagen (Chen and Xu, 2015).

**Table 3.**
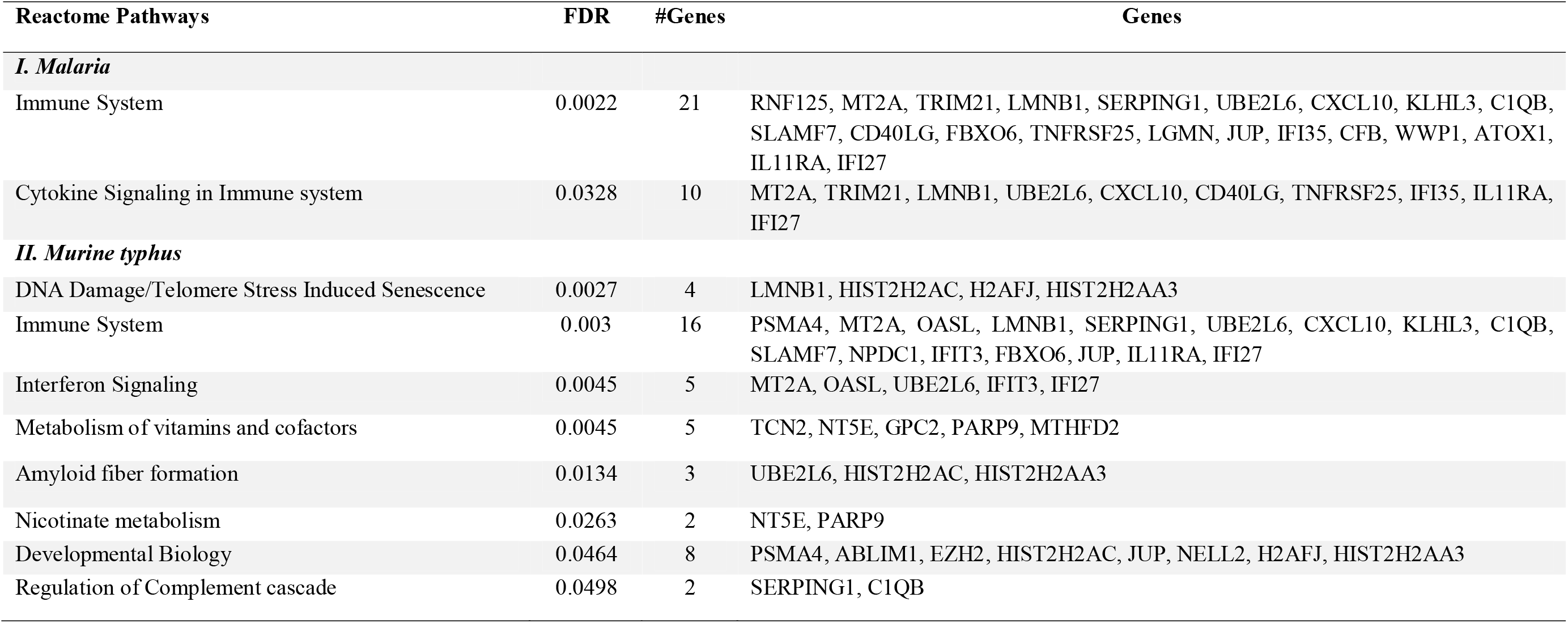
Enriched reactome pathways for DEGs in malaria and murine typhus.

The MT regulatory gene network (Figure 2D) displayed the involvement of a total 35 comprising 19 upregulated and 16 downregulated genes with log2FC ranging from 1.02326 (for LAP3) to 2.46668 in (for IFI27), and from -1.02326 (for C9 or f123) to -2.46668 (for LRRN3), respectively. The immune system and DNA damage/telomere stress induced senescence were the most significant reactome pathways in MT group associated with genes including PSMA4, MT2A, OASL, LMNB1, SERPING1, UBE2L6, CXCL10, KLHL3, C1QB, SLAMF7, NPDC1, IFIT3, FBXO6, JUP, IL11RA, IFI27, HIST2H2AC, H2AFJ, and HIST2H2AA3 (Table 3).

The over and under stimulated genes among the ST, DG, ML and MT disease groups are displayed in the heat map (Figure 3A), which indicated the presence of 67 DEGs with highly overexpressed and under stimulated genes in DG in the upper and lower respective panels, while 3 genes including IL32, LOC729530, PLTP were highly induced in ST. The number of unique signature genes for ST, DG, ML and MT were 31, 267, 4, and 3 respectively, while the number of overlapping genes in the ST group with the DG, ML and MT were 114, 42, and 54, respectively; 32 genes belonged commonly to all the four categories, as represented by the Venn diagram (Figure 3B). The ST category consisted of 530 PPIs (Figure 3C), with an enrichment of 10^−16^. The most interacting proteins within this network were related to immune response including T-lymphocyte differentiation antigen T8/Leu-2 (CD8A) and T-cell surface glycoprotein CD8 beta chain (CD8B), T-cell surface glycoprotein CD3 gamma chain (CD3G), small and large ribosomal subunits RPS23 and RPL22 with interaction score 0.961 – 0.999. Among the enriched reactome pathway, interferon signaling involving 22 genes namely USP18, OAS3, EIF2AK2, TRIM21, OASL, HERC5, RPS27A, UBE2L6, STAT2, MX2, OAS2, STAT1, IFIT1, IFIT3, IFIT2, ISG15, DDX58, RSAD2, IRF7, MX1, OAS1, and IFI35 were the key players in the pathogenesis of currently described four disease types (Figure 3D and Table 4). Defense response to virus was the most significantly expressed biological process common to these diseases, associated with 22 genes including OAS3, EIF2AK2, OASL, RTP4, IFIH1, HERC5, CXCL10, STAT2, MX2, OAS2, STAT1, IFI44L, IFIT1, IFIT3, IFIT2, ISG15, DDX58, TLR7, RSAD2, IRF7, MX1, OAS1 (Table 4).

**Table 4.**
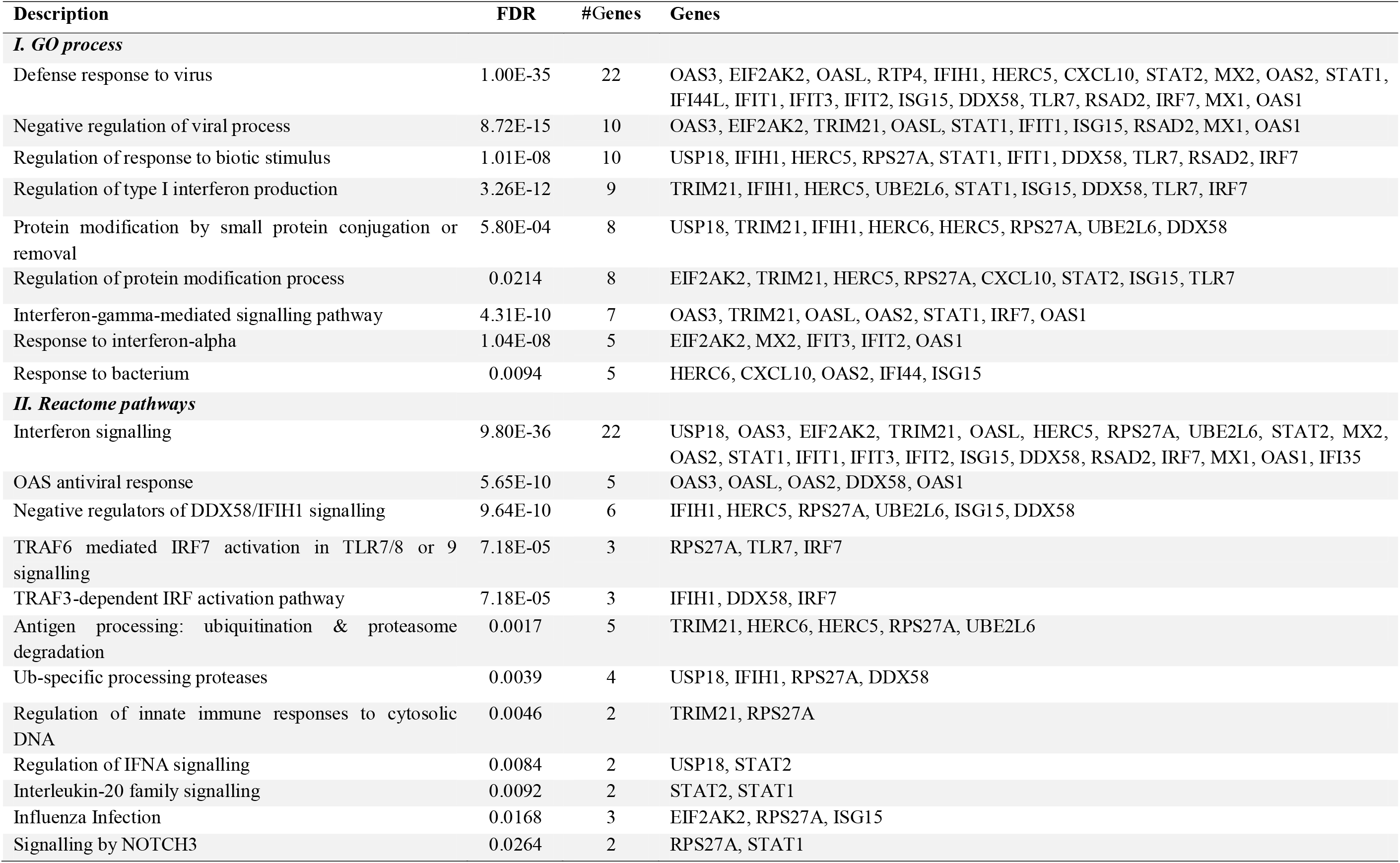
Hub genes of ST, DG, ML and MT disease groups.

**Figure 3:**
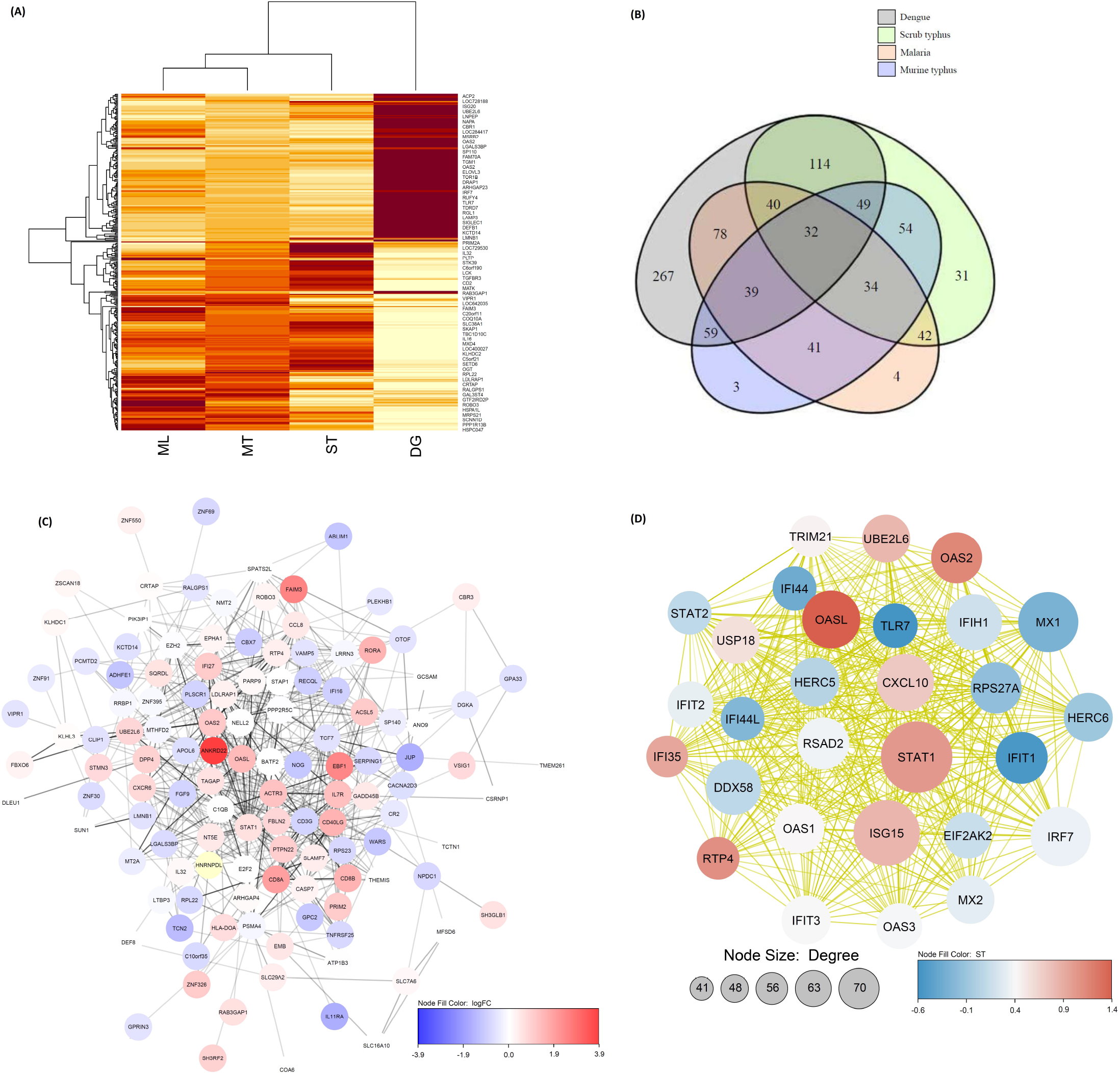
(A) Heatmap of differentially expressed genes among the ST, DG, ML and MT disease groups compared to health controls; (B) Venn diagram showing number of genes belonging to ST, DG, MT and ML disease groups; (C) protein-protein interaction (PPI) in ST; (D) top ranked 29 hub genes in ST, DG, MT and ML disease groups by maximal clique centrality.

The DG disease category comprised of 1766 PPIs (Figure 4A), with an enrichment of 10^−16^, the most interacting proteins within this network included small and large ribosomal subunits involved in protein synthesis (RPS23, RPS27A, RPL13A, RPL22, RPL37) with interaction score 0.999. The MT disease category demonstrated 47 PPIs with an enrichment of 5.89 × 10^−9^ (Figure 4B); the most interacting proteins within this network included core components of nucleosome H2A histone family member J (H2AFJ), histone cluster 2 (HIST2H2AC and HIST2H2AA3), interferon-induced protein with tetratricopeptide repeats 3 as an inhibitor of cellular as well as viral processes (IFIT3), interferon alpha-inducible protein 27 (IFI27), 59 kDa 2’-5’-oligoadenylate synthase-like protein (OASL), serpin peptidase inhibitor (SERPING1), complement component 1 (C1QB), with interaction score 0.961 – 0.995. The ML disease expressed 50 PPIs with an enrichment of 2.36 × 10^−16^ (Figure 4C); the interaction score ranged from 0.96 to 0.979 for the mostly ML-associated proteins including mitochondrial glycine degrading aminomethyltransferase (AMT), bifunctional methylenetetrahydrofolate dehydrogenase/cyclohydrolase (MTHFD2), SERPING1, C1QB, HIST2H2AC, HIST2H2AA3, interferon-induced 35 kDa protein (IFI35), IFI27.

**Figure 4:**
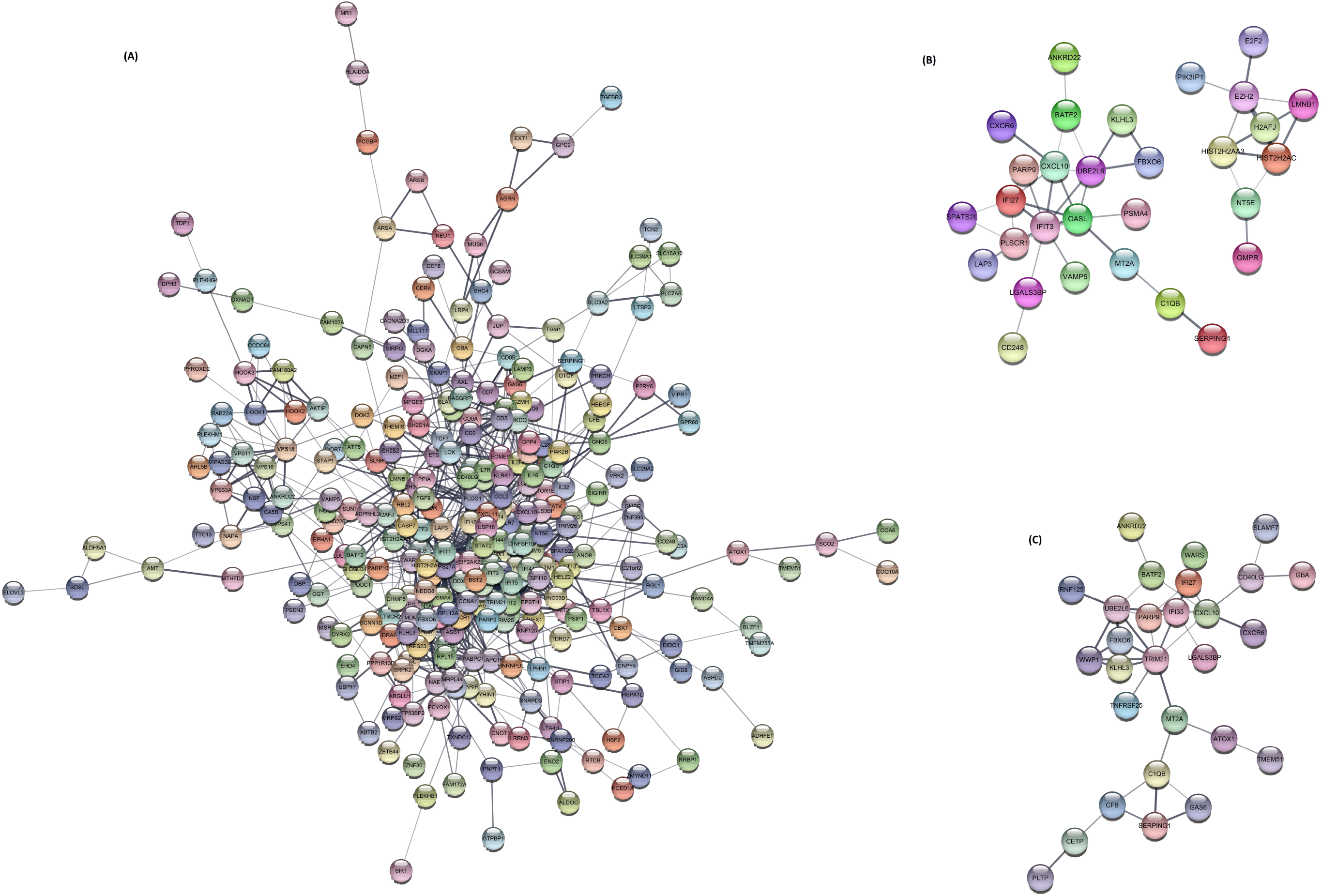
protein-protein interaction (PPI) in (A) dengue, (B) murine typhus, (C) malaria.

Bacterial organisms stimulated the expression of a large panel of genes including type I interferon, interferon-stimulated, inflammatory, apoptosis-related genes and induced an M1-type gene response in macrophages, accounting for the local and systemic inflammation observed in ST, and that interferon-gamma may be useful as an adjuvant treatment of patients with scrub typhus. (Tantibhedhyangkul et al. 2013). The complement system as a link between the adaptive and innate immune system, was implicated in viral antagonism and immune evasion; human proteins linked to the complement and coagulation cascade, the centrosome, and the cytoskeleton were enriched among the dengue viral interaction partners (Khadka et al. 2011). In ST, coagulation activation was prominent and linked to a strong proinflammatory response with high level of inflammatory cytokines expression (except IL12) compared to MT cases that displayed expression of high level of endothelium-derived markers, as demonstrated in the early phase of infection with *R. typhi* and *O. tsutsugamushi* contributing to disease differentiation (Paris et al., 2012).

Comparison of 635 gene sets screened from RNA microarrays of ST, DG, ML, MT cases, and healthy controls suggested that a subset of 31 genes were found unique to the ST group, which could be used as specific signature of scrub typhus significantly overrepresenting translocation of ZAP-70 to immunological synapse, and phosphorylation of CD3 and TCR zeta chains, involving PTPN22 and CD3G genes. The representative biomarkers of *O. tsutsugamushi* infection found in the current study will help in differential diagnosis of ST mimicking other acute febrile illnesses. These genes could be useful for investigating the molecular pathophysiology of ST and for discovering novel drug targets and vaccine developments.

## Data Availability

The data analyzed in the current study are publicly available as mentioned in the manuscript, and new data generated are retained by the authors of tis paper.

## Author’s contribution

Both MM and SM designed the study, analysed and interpreted data, discussed and wrote the manuscript.

